# The impact of an ageing population on pandemic risk in England

**DOI:** 10.64898/2026.09.11.26362845

**Authors:** Thomas Rawson, Pablo N. Perez-Guzman, Edward S. Knock, Christian Morgenstern, Daniel Valdenegro, Katy A.M. Gaythorpe, Lilith K. Whittles, Wes Hinsley, Raphael Sonabend, Richard G. FitzJohn, Anne Cori, Jennifer Beam Dowd, Jakub Bijak, Neil M. Ferguson, Melinda C. Mills

## Abstract

**Background:** England’s population is ageing rapidly, with those aged 80 years and older projected to increase 85% by 2047. Aging populations may change the dynamics of future pandemics, since older individuals face higher risks of severe outcomes and require a disproportionate share of early vaccine supply, for certain respiratory infections, despite having fewer social contacts. The net pandemic-risk implications of these competing effects cannot be resolved without dynamic transmission modelling.

**Methods:** We used an age-structured stochastic SARS-CoV-2 transmission model, previously calibrated to the realtime COVID-19 epidemic across seven NHS regions of England. Running the fitted model under the ONS principal population projection for 2047 as a counterfactual, we held all non-pharmaceutical interventions fixed and scaled vaccine supply to overall population growth. Nine counterfactual scenarios spanned three vaccine deployment strategies and three ONS migration assumptions.

**Findings:** The basic reproduction number remained largely unchanged by the projected age distribution. However, under the central counterfactual scenario, cumulative hospital admissions in the first two years of the pandemic were 674,004 - a 40·3% increase over the historic baseline - and hospital deaths increased by 49·4%, disproportionate to the 13·9% total population increase. Trajectories were similar before the Delta variant emerged but diverged thereafter, as a larger elderly cohort delayed vaccine protection in younger, higher-contact groups. Projected burden was robust to migration assumptions (664,451–689,833 admissions). Vaccine deployment strategy had substantially greater influence, with admissions ranging from 625,859 (optimistic age-scaled deployment) to 1,193,883 (non-scaled).

**Interpretations:** Population ageing will disproportionately amplify England’s pandemic burden, driven primarily by the interaction between a growing elderly cohort and fixed age-ordered vaccine allocation, rather than by population growth alone. This vulnerability is largely independent of future migration uncertainty but is strongly modifiable through demographically-adaptive vaccine deployment planning developed before, not during, the next pandemic.

**Funding:** Moh Family Foundation; UK Medical Research Council; Leverhulme Trust.

**Research in context:** *Evidence before this study:* We searched PubMed up to June 19th, 2026, with no language restrictions using the following search terms: (COVID-19 OR SARS-CoV-2 OR pandemic OR “infectious disease*”) AND (“population ageing” OR “population aging” OR “demographic ageing” OR “demographic aging” OR “ageing population” OR “aging population” OR “demographic change” OR “demographic projection*”) AND (model* OR simulat* OR “transmission dynamics”). We found five studies modelling the effect of demographic change on infectious disease transmission. However, none of these studies were fitted to the real-world trajectory of a historical pandemic, nor combined this with an official national population projection. Additionally, none of the studies considered non-pharmaceutical interventions or vaccine roll-out policy, to isolate the specific contribution of population ageing to pandemic risk in England or elsewhere.

*Added value of this study:* We are the first to combine an age-stratified pandemic model, previously validated against England’s real COVID-19 trajectory, with an official long-range demographic projection and a previously set vaccine deployment policy, isolating the effect of demographic change from policy response. We show that an ageing population substantially worsens pandemic outcomes not simply due to age-adjusted risk of severe disease, but by straining age-prioritised vaccine allocation — a mechanism not previously identified. Different migration scenarios, despite shifting the population size projected for 2047 by ± five million people, had little effect, confirming age structure as the key driver. These findings have direct relevance for pandemic preparedness planning and vaccine allocation strategy in ageing societies.

*Implications of all the available evidence:* Our study shows that, even with no change to non-pharmaceutical interventions, the same pandemic as COVID-19 could result in a substantially greater burden in an older future population, driven less by the larger number of elderly people than by the strain ageing places on age-prioritised vaccine allocation. Pandemic preparedness plans built around current demographics instead of projected ones therefore underestimate future risk, and vaccine allocation strategies optimised for today’s population structure will need revisiting as populations continue to age. Future work should test whether alternative allocation strategies can mitigate this effect, and extend this approach to household-structured models and to other ageing populations beyond England.

## Introduction

Europe is undergoing an unprecedented demographic shift. Increasing lifespans and decreasing birth rates mean the proportion of the EU’s population aged 80 years and above is projected to increase from 6·2% in 2025, to 15·3% by 2100.^1^ For England, the Office for National Statistics (ONS) projects that the population will grow from 57·1 million in 2022 to 65·4 million by 2047, driven almost entirely by net migration, while the number of people of pensionable age will increase by 25·5%, faster than any other age group.^2^ Alongside this, the Office for Budget Responsibility (OBR) forecasts a rapidly increasing old-age dependency ratio (the proportion of people above state pension age, compared to the working age population below it), from 31% in 2023 to 47% by 2074.^3^ England’s Chief Medical Officer, in their 2023 annual report on health in an ageing society, called for research into multimorbidity, frailty, and social care to be accelerated, while specifically highlighting “too little investment into infection in older adults” to date.^4^

The implications of population ageing for chronic, non-communicable disease are comparatively well understood, and largely intuitive: as more people survive into older age, more people will live with frailty and accumulating multimorbidity, increasing demand on health services.^5^ The implications of population ageing for infectious disease burden are less straightforward. At the individual level, older individuals face worse outcomes from communicable diseases, with markedly higher rates of hospitalisation observed for, among others, seasonal influenza,^6^ respiratory syncytial virus,^7^ and COVID-19.^8^ However, at the population level, the impact of the population’s age distribution is more complicated. Older age groups have repeatedly been observed to have a lower average number of daily contacts,^9,10^ a trend that was observed throughout all stages of non-pharmaceutical intervention during the COVID-19 pandemic in England.^11^ This effect means that a proportionately older population would display a lower force of infection (and reproduction number) than a younger population of the same size. The epidemiological impacts become further complicated once the logistics of vaccine deployment are considered. Emergency vaccination campaigns are typically prioritised by age and clinical risk,^12^ meaning an older population would require a disproportionately larger share of any fixed vaccine supply before younger groups could be reached. Understanding how these factors mechanistically combine to shape future healthcare demand requires explicit dynamic transmission modelling.

The COVID-19 pandemic generated an unprecedented depth of age-stratified epidemiological data, offering a rare empirical opportunity to study how demographic change influences the spread of disease. In this study, we use an age-structured transmission model previously calibrated against the real trajectory of the COVID-19 pandemic in England, and used to inform UK pandemic policy, to ask how the pandemic risk landscape of England would change under the ONS principal projection of its 2047 population. By recreating the non-pharmaceutical interventions and the Joint Committee on Vaccination and Immunisation (JCVI)’s age-prioritised vaccine roll-out actually used during the pandemic as a counterfactual, we isolate the contribution of future population ageing to pandemic risk from the contribution of policy response.

## Methods

### Epidemiological model and fitting

We use a previously described^13,14,15,16^ stochastic, age- and region-structured compartmental model of SARSCoV-2 transmission in England. The model has a susceptible-exposed-infectious-removed (SEIR) structure, stratified into 17 age classes (5-year bands {0-4, 5-9, …, 75-79} and a final 80+ years class) and the seven National Health Service (NHS) England regions, with explicit clinical pathways for community and hospital care, including general beds and intensive care.

Heterogeneous contact rates between age groups were informed by the POLYMOD contact matrix for England,^9^ modified by a piecewise-linear, time-varying transmission multiplier fitted to capture the impact of non-pharmaceutical interventions, school holidays, and other behaviourally-relevant events. Age-specific probabilities of hospitalisation, intensive care admission, and death were modelled using fitted age-splines, further adjusted by time-varying multipliers capturing changes in clinical practice and healthcare-seeking behaviour over the course of the pandemic.

The model accounts for sequential strain replacement between wildtype, Alpha, Delta, and Omicron BA.1, with variant-specific transmissibility and severity informed by regional variant frequency data, and partial cross-immunity between variants informed by the literature. Vaccination is modelled across seven classes (unvaccinated; first dose, before and after full effectiveness; second dose; waned second-dose protection; booster dose; and waned booster protection), with age- and vaccine type-specific effectiveness against infection, symptomatic disease, severe disease, death, and onward transmission. The number, platform, and age of vaccine recipients by region and date were informed by NHS England administration data, implicitly capturing the real-world prioritisation of doses by age and clinical risk group, as recommended by the JCVI.^12^ Full model equations, parameters, and data sources are described in Sections 1-4 of the Supplementary Appendix.

The model was fitted in a Bayesian evidence synthesis framework via particle Markov Chain Monte Carlo to multiple regional epidemiological data streams between March 16, 2020 and February 24, 2022, including community PCR positivity (Pillar 2), the Office for National Statistics COVID-19 Infection Survey, the REal-time Assessment of Community Transmission (REACT) study, hospital admissions, community and hospital deaths, and general and intensive care bed occupancy. We use the model fit as presented in Perez-Guzman et al. (2023)^13^ as our baseline factual scenario, compared against counterfactuals for different population and vaccine allocation scenarios. All previous model fit plots are provided in Section 6 of the Supplementary Appendix.

### ONS population projections and demographic scenarios

The baseline scenario used the 2020 mid-year ONS estimates for age-specific population counts in each of the seven NHS regions.^17^ For our central counterfactual, we instead consider the latest ONS “migration category variant” projections (at the NHS region level) for 2047^18^ - the furthest the ONS currently projects to at this geographic level. The ONS specifically recommends this dataset as the principal projection to consider, due to its use of data on migration and resident population from across several administrative records.^19^ The ONS also provide additional projections for varying assumptions on future migration trends, for which we also consider the ‘low’ and ‘high’ international migration scenarios.^20,21^ Under the different scenarios, net migration for the whole of the UK is assumed to stabilise at 340,000 persons per year in the central “migration category variant”, 120,000 in the ‘low’ scenario, and 525,000 in the ‘high’ scenario. These different migration scenarios do not broadly alter the age composition of the population, but rather the total number (Figure S10 of the Supplementary Appendix). We present the baseline and central counterfactual populations in Figure 1. Alternative scenario-based populations are provided in Section 5.1 of the Supplementary Appendix.

**Figure 1.**
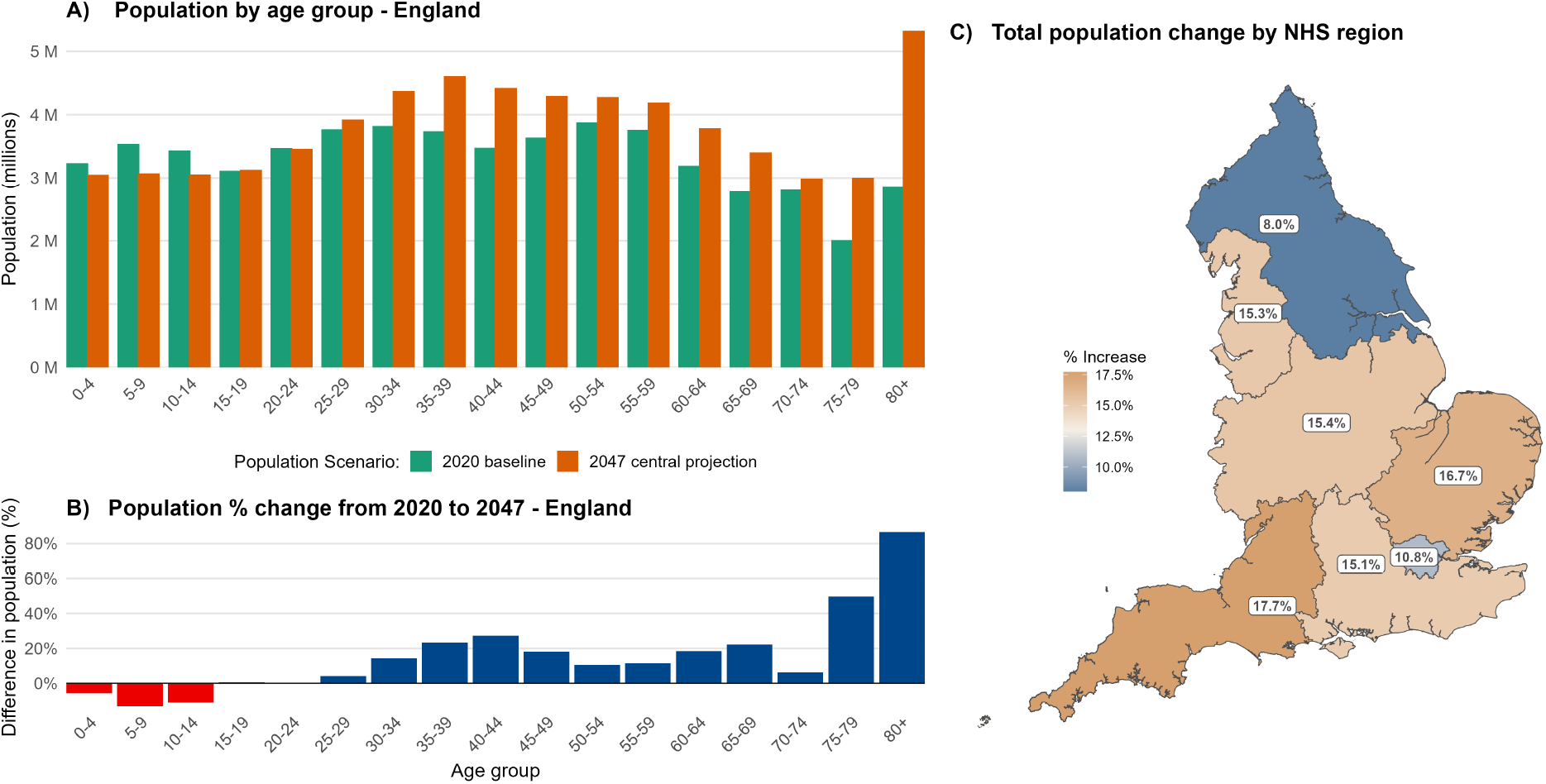
**A)** Population of England by age-group: “2020 baseline” shows the ONS mid-2020 estimates,^17^ and “2047 central projection” refers to the ONS baseline variant projection for that year.^18^ **B)** Percentage difference between the age-group populations of the baseline and central counterfactual. Red denotes age groups with a reduced total population, blue with an increased population. **C)** Total population increase by 2047 in each of the seven NHS regions of England. Region-specific versions of **(B)** are provided in Section 5.1 of the Supplementary Appendix.

Age-specific contact rates for each population scenario were derived from the POLYMOD social contact survey for the United Kingdom,^9^ reweighted via the socialmixr R package^22^ to ensure reciprocity of total contact events between age groups under each scenario’s population structure (Figure 2A-B). Fewer contacts occur with younger individuals, and more contacts occur with older individuals (Figure 2C), reflecting the projected changes to the respective population sizes of each age group. The difference between the respective transmission matrices is shown in Figure 2D. Minimal change is seen across the majority of age-pairings, as increases in raw contact counts under an enlarged older population are largely offset once expressed on a per-individual basis.

**Figure 2.**
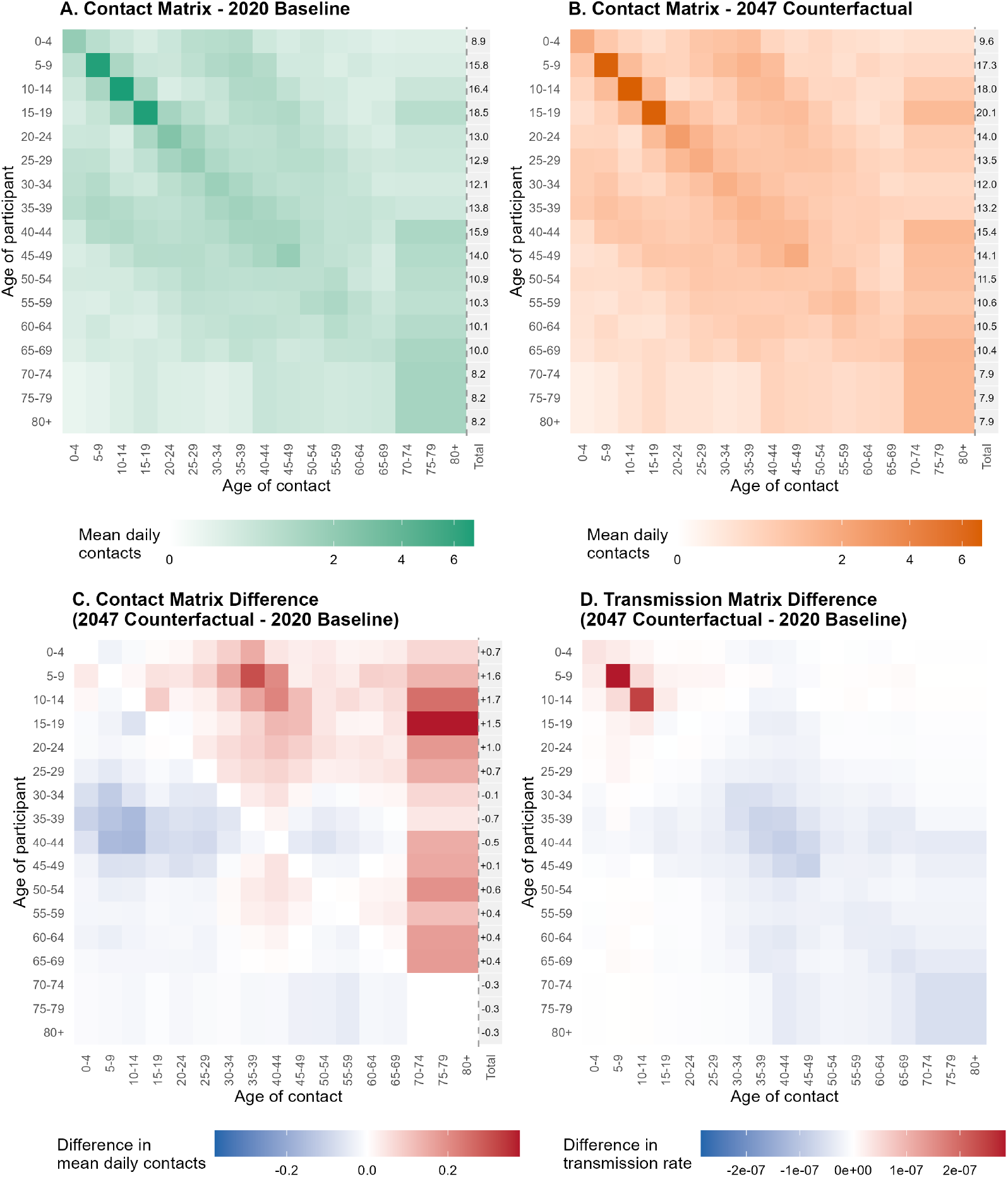
Contact and transmission matrix structure under historic baseline (2020) and central counterfactual (2047) population projections for England. **A, B)** Estimated mean number of daily contacts by age of participant (rows) and age of contact (columns), derived from the POLYMOD contact survey,^9^ using the socialmixr R package,^22^ and reweighted for reciprocity using the baseline and counterfactual population age structures respectively. The rightmost column gives each age group’s total mean daily contacts, summed across all contact ages. **C)** Difference in mean daily contacts (counterfactual minus baseline). Positive (red) values indicate more contacts under the 2047 population structure; the total column gives the net change in each age group’s total daily contacts. **D)** Difference in the corresponding per capita transmission rate (mean daily contacts divided by the population size of the contact age group). We assume identical contacts for the 70-74, 75-79, and 80+ age groups, as POLYMOD data only extends as far as a 70+ age category.

### Counterfactual vaccine deployment strategies

Real-world vaccine administration in England, by NHS region, age group, and date, was used to inform the baseline model as previously described.^13^ However, one can argue that the historical record of daily doses administered cannot be applied unaltered to a larger future population: it reflects the manufacturing and logistical capacity required to vaccinate the population as it existed at the time, whereas the central ONS projections for 2047 point towards a 13·9% total population increase. We therefore consider three assumptions about how vaccine deployment might be expected to change alongside population growth.

In the first, most conservative scenario, the historical number of doses administered each day, by region, age, and type, was applied directly to the counterfactual population, unadjusted for population size (“historic vaccinations”). This represents a fixed absolute vaccine supply, implying no expansion in deployment capacity despite a larger population in need of protection. We show this scenario in Figure 3A, where the final percentage point differences reflects the percentage change in population, as shown in Figure 1B.

**Figure 3.**
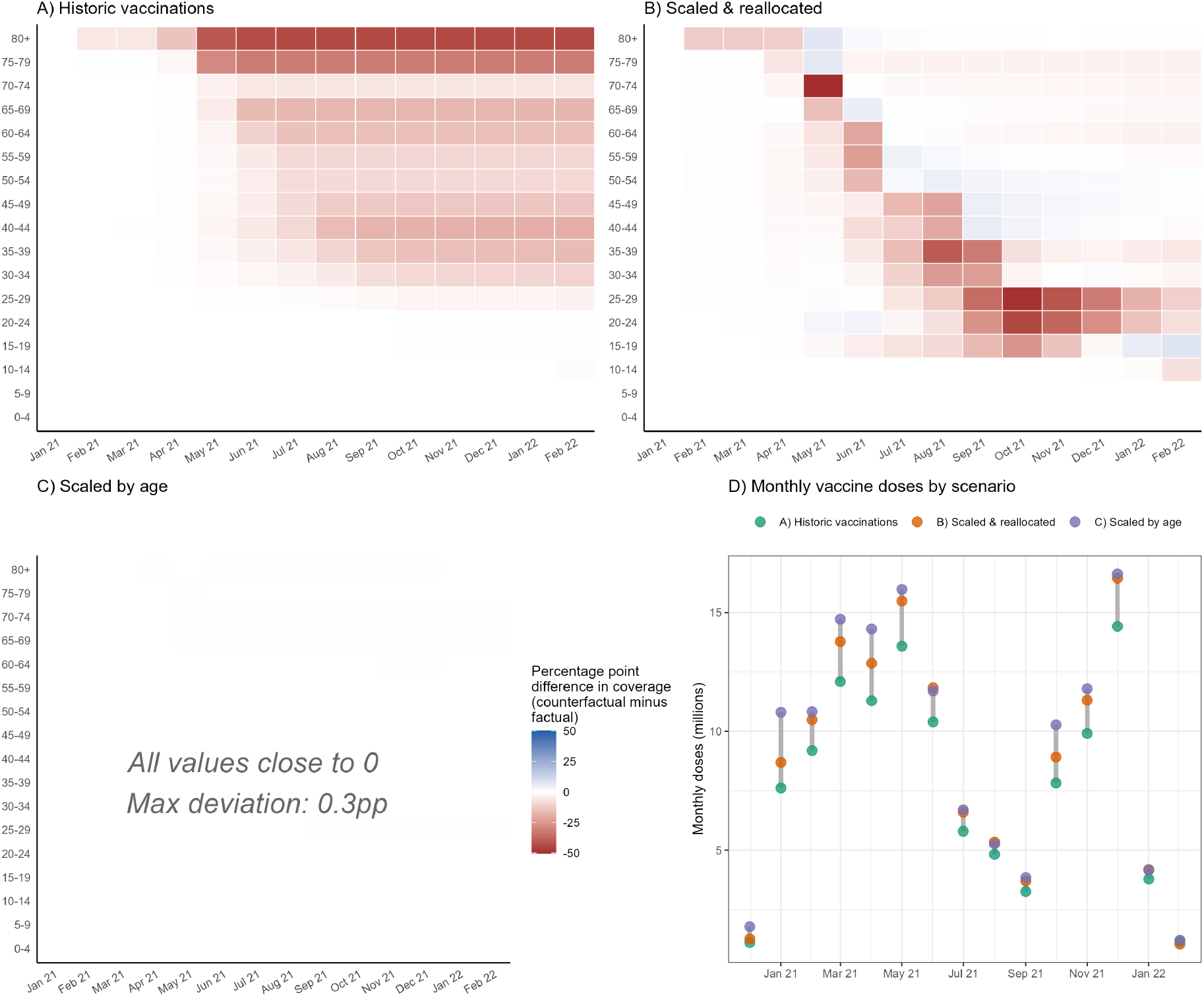
**A-C)** Difference in cumulative vaccine coverage by age group and month, between the factual pandemic and three counterfactual vaccine deployment scenarios, under the central 2047 population projection for England. Coverage is defined as the cumulative proportion of each age group that has completed the primary vaccination course (i.e. reached the “Full dose 2 protection” stratum). Colour denotes the percentage point difference in coverage between scenarios (counterfactual minus factual) at each age group and month; red indicates lower proportional coverage under the counterfactual relative to the factual rollout, blue indicates higher proportional coverage. **A)** “Historic vaccinations”: the historical number of doses administered each day is applied, unscaled, to the larger counterfactual population. **B)** “Scaled & reallocated” (our central vaccine deployment assumption): total daily doses are scaled by each region’s overall population growth and reallocated following JCVI-recommended priority order. **C)** “Scaled by age”: doses administered to each age group are instead scaled by that age group’s own population growth, independent of all other age groups; maximum absolute deviation from the factual rollout under this scenario is annotated. **D)** The total new vaccines scheduled to be administered every month under the three vaccine scenarios assuming the central population projections.

In our central vaccine scenario, the total number of doses administered each day in each NHS region was instead scaled by that region’s overall population growth factor under the relevant demographic scenario, and the resulting (larger) daily supply was then re-allocated across age and clinical risk groups following the same priority order recommended by the JCVI and observed in the historical roll-out^12^ (“scaled and reallocated”). Under this assumption, the oldest and most clinically vulnerable groups continue to be vaccinated first. However, because the daily supply of vaccines is scaled only to the population’s overall total growth, rather than to the disproportionately larger growth of the older age groups specifically, this larger older-age population takes correspondingly longer to be fully protected than it did historically - in turn delaying the point at which younger age groups begin receiving vaccine doses. We show this scenario in Figure 3B, where the strong diagonal reflects the successive age groups being delayed in receiving their doses, due to the extended time needed to vaccinate the older age groups.

In a third, more optimistic scenario, the historical number of doses administered each day to each specific age group was instead scaled by that age group’s own population growth factor, independent of all other age groups (“scaled by age”). Under this assumption, vaccine supply expands in lockstep with the demographic change occurring within each age group, such that, for example, the unprecedented growth in the 80 years and older population is matched by a proportionate increase in the doses available to that group on the equivalent day of the historical roll-out. This represents an optimistic upper bound on deployment capacity, as it implicitly assumes manufacturing and distribution networks capable of dynamically tracking age-specific demand, rather than a fixed regional supply subsequently reallocated by priority order. We show this scenario in Figure 3C, where no variation in percentage point difference is observed - this scenario assumes we match the proportional increase in population with vaccination deployment.

Additional figures demonstrating the difference in vaccination rollout are provided in Section 5.2 of the Supplementary Appendix. These three vaccine deployment assumptions were each run under the low, central, and high migration population scenarios described above, producing nine counterfactual scenarios in total. The central vaccine deployment and central migration scenario forms our primary counterfactual; the remaining eight combinations are presented as sensitivity analyses.

### Outcome measures and scenario simulation

For each demographic and vaccine deployment scenario, we ran the fitted transmission model forward from the same posterior parameter distribution used to reconstruct the historical epidemic in England, holding the timing and strength of all non-pharmaceutical interventions fixed at their fitted historical values. This ensures that any differences between the baseline and counterfactual scenarios are attributable to the demographic and vaccine deployment assumptions under consideration, rather than to differences in fitted transmission, severity, or behavioural parameters. All scenarios were simulated over the period encompassed by the original model fit, from March 16, 2020 to February 24, 2022.^13^ All scenarios were run with 50,000 draws from the underlying posterior distributions and subsequent stochastic model runs. All outcomes are summarised as the mean and 95% credible interval across stochastic model realisations and posterior parameter draws.

## Results

We plot the key epidemiological differences between the historic baseline and our central counterfactual scenario in Figure 4. The central 2047 counterfactual analysis sees a mean cumulative total of 674,004 hospital admissions and 164,977 deaths in hospital within the simulated time period. This is an increase of 40·3% and 49·4% respectively from the historic baseline values. We see that in the period before the emergence of the Delta variant on March 8th 2021, the dynamic behaviour of the epidemiological trajectories are broadly similar between the two scenarios (panels 4A and 4C), differing only in peak magnitude. However, after the emergence of Delta (and critically, after substantial rollout of the vaccination campaign) the epidemiological patterns begin to diverge. The central counterfactual demonstrates two clear additional peaks in hospital admissions and deaths around August 2021 and January 2022. While the infection hospitalisation ratio (IHR; panel 4E) and infection fatality ratio (IFR, panel 4F) can be seen to diverge between the two scenarios to a small degree in these periods, the divergence is not enough to fully explain the near doubling of admissions and deaths around these peaks. We reproduce Figure 4 for additional counterfactual sensitivities in Section 5.3 of the Supplementary Appendix.

**Figure 4.**
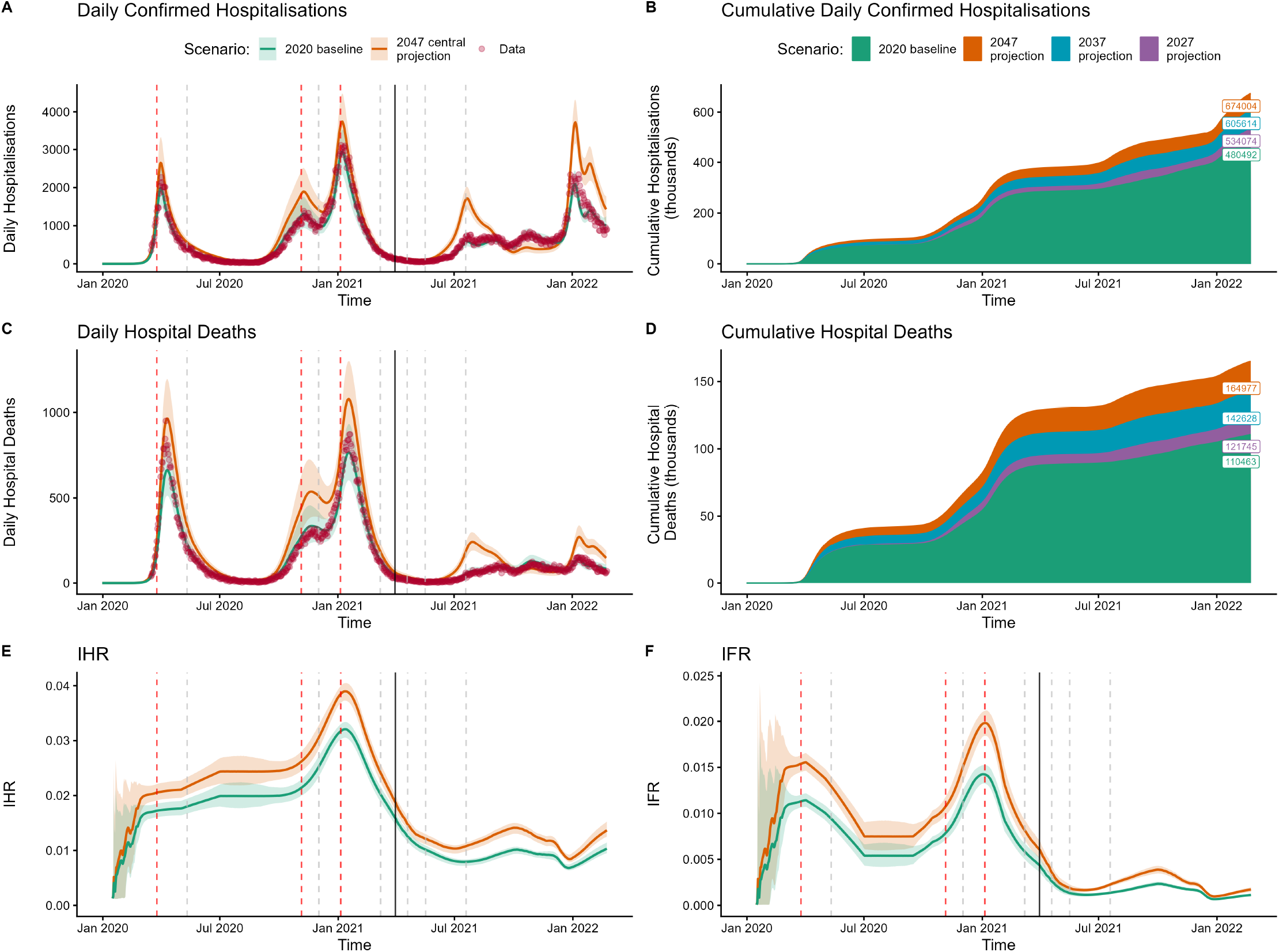
Epidemiological differences between the historic baseline and counterfactual simulations in England. Panels **A)** and **C)** plot daily confirmed hospital admissions and deaths in hospital with COVID-19, respectively, under the baseline and central 2047 counterfactual only; red points show the underlying historic data. Panels **B)** and **D)** plot the cumulative totals of the same measures under the baseline and all three counterfactual population horizons (2027, 2037, and 2047), with text overlays giving the final respective totals on 24 February 2022. Panel **E)** plots the time-varying infection hospitalisation ratio (IHR) – the proportion of all infections resulting in hospitalisation – and panel **F)** the time-varying infection fatality ratio (IFR) – the proportion of all infections resulting in death, in hospital or the community – under the baseline and central 2047 counterfactual. Throughout, green shows the baseline model fit; orange, blue, and purple show the 2047, 2037, and 2027 counterfactual population projections, respectively, where shown. Red dashed vertical lines signify points at which stay-at-home ‘lockdown’ restrictions were enacted, and grey dashed vertical lines represent points where restrictions were ended, or successively eased in the case of the third lockdown.^23^ The solid black vertical line denotes the point at which the Delta variant emerged. All counterfactuals assume the central migration assumption and the central “scaled and reallocated” vaccine deployment scenario.

We explore the mechanisms driving the epidemiological differences in Figure 5. Panel 5A shows that the number of daily new symptomatic cases is very similar between the two scenarios before the emergence of Delta (solid black line), with the mean values of both scenarios within the 95% credible intervals of the other. This is shown more explicitly in panel 5B, plotting the effective reproduction number, 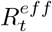 - the average number of secondary infections at time *t* accounting for immunity (infection- and vaccine-derived) and NPI policies present at that time in the population. 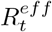 is almost identical between the two scenarios before the emergence of Delta. 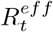 initialises at 2·58 (2·47-2·73 95% CrI) in the baseline, and 2·60 (2·48-2·74 95% CrI) in the counterfactual of 5B. The trajectories diverge however from approximately June 1st 2021. The post-Delta divergence in epidemic trajectory can therefore only be explained by the one factor that dynamically differs between the two scenarios - namely, the vaccination deployment.

**Figure 5.**
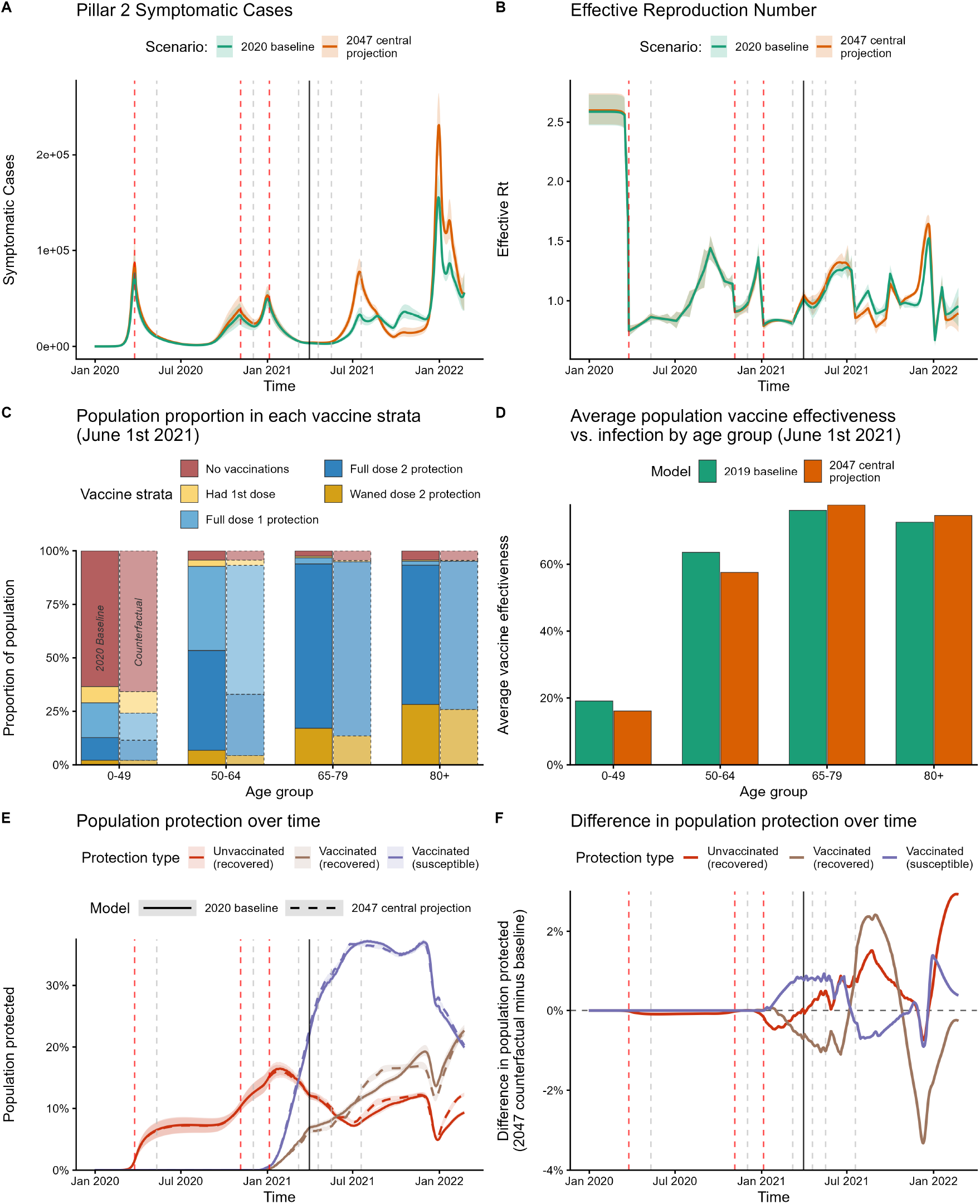
Differences in daily cases, vaccination, and protection against infection between the historic baseline and counterfactual simulations in England. **A)** The daily number of new pillar 2 symptomatic cases between the historic baseline and central counterfactual. **B)** The associated effective reproduction number 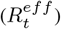 over time. **C)** The proportions of the population split across vaccine strata for four different age groups. The solid colours depict the historic baseline and the faded colours in dashed lines depict the counterfactual. Vaccine effectiveness values for each strata are given in section 3 of the supplementary appendix. **D)** shows how the vaccine delivery shown in panel **C)** translates to average protection against infection for each of the four age groups. **E)** The proportion of the population considered immune to infection over time. Red denotes acquired immunity from prior infection, purple denotes immunity from vaccination and no prior infections, brown denotes protection from both vaccination and prior infection. Dashed line denotes the counterfactual and solid line the historic baseline. **F)** The absolute difference between the mechanisms of protection, the dashed line minus the solid line of panel **E).** In panels **A), B), E)** and **F)** red dashed vertical lines signify points at which stay-at-home ‘lockdown’ restrictions were enacted, and grey dashed vertical lines represent points where restrictions were ended, or successively eased in the case of the third lockdown.^23^ The solid black vertical line denotes the point at which the Delta variant emerged. All counterfactuals assume the central migration assumption and the central “scaled and reallocated” vaccine deployment scenario. Shaded regions denote 95% credible intervals.

Due to the greater population of older individuals in the central counterfactual, it takes a longer period of time to vaccinate these cohorts, meaning the younger age groups go longer without vaccine protection. Panel 5C shows this by plotting the proportion of four aggregated age groups within each vaccine class on June 1st 2021, when the trajectories begin to diverge. While vaccination levels are similar in the 65-79 and 80+ categories, we see that administration of doses to the younger categories is lagging behind in the counterfactual, with substantially fewer second doses administered. This directly translates into reduced protection against infection in the younger age cohorts (panel 5D), causing an increase in infections. This in turn, increases the overall force of infection on the entire population.

Panel 5E shows how overall population protection against infection varies over time between the two scenarios, and whether they are protected by vaccination, previous infection, or both. Protection from acquired immunity is identical throughout 2020, and the divergence in July 2021 is driven by reduced vaccination levels in the previouslyinfected. This increase in cases then results in overall protection from previous infection greatly increasing in the counterfactual, driving 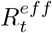 below 1 (panel 5B). This imbalance is then later offset by the immune escape features of the Omicron variant in late 2021. Further highlighting the role of vaccines in driving this epidemiological difference, the time-varying basic reproduction numbers of both scenarios, removing the role of immunity in the population, remain within one another’s 95% credible intervals across the entire time series (Section 5.4 of the Supplementary Appendix).

We assess the sensitivity of overall hospital burden to alternative migration and vaccine deployment assumptions in Figure 6. Varying the ONS migration scenario had only a minor effect on projected cumulative admissions under the central vaccine deployment assumption: under the 2047 counterfactual, cumulative admissions ranged from 664,451 (low migration) to 689,833 (high migration), a span of less than 4% around the central estimate of 674,004. A small difference considering the total population is 4,179,341 fewer in the low migration scenario, and 5,360,544 greater in the high migration scenario. These migration scenarios vary in total population size, but are broadly similar in age composition (Figure S10 of the Supplementary Appendix). Migration uncertainty was similarly inconsequential for the 2027 and 2037 intermediate projections. In contrast, vaccine deployment strategy had a substantially larger influence on outcomes than migration uncertainty.

**Figure 6.**
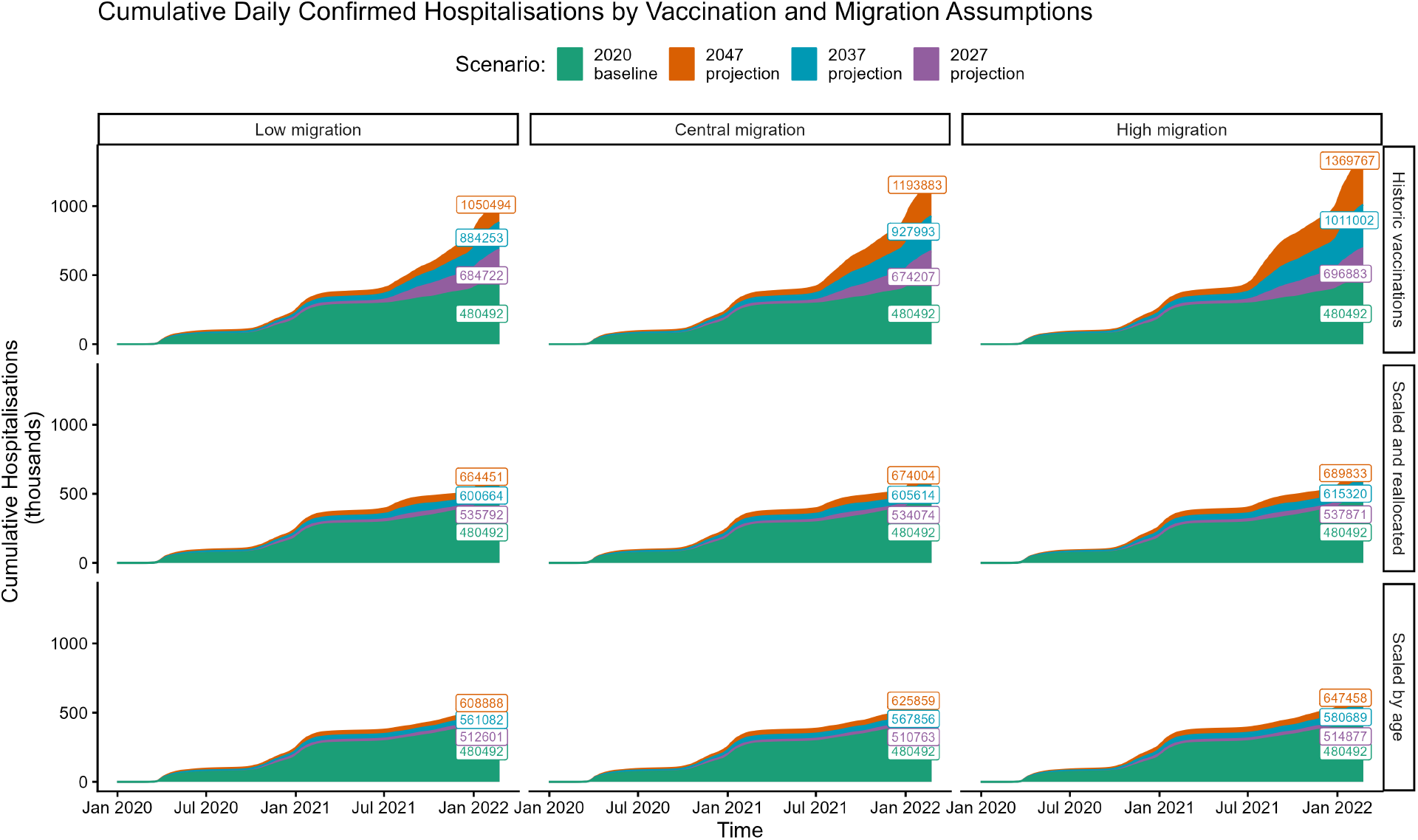
Sensitivity of cumulative confirmed hospital admissions to vaccine deployment strategy and migration assumptions. Each panel plots cumulative daily confirmed hospital admissions over the study period for the historic baseline (green) and three counterfactual population horizons - 2027 (purple), 2037 (blue), and 2047 (orange) - with text overlays giving the final cumulative total for each scenario on 24 February 2022. Columns correspond to the three ONS migration variant assumptions (low, central, and high international migration), and rows correspond to the three vaccine deployment strategies described in the Methods: **historic vaccinations**, in which the historical absolute number of doses administered is applied unscaled to the larger counterfactual population; **scaled and reallocated** (our central assumption), in which total daily doses are scaled by each NHS region’s overall population growth and reallocated following JCVI-recommended age-priority order; and **scaled by age**, in which doses administered to each age group are scaled by that age group’s own population growth factor, independent of all other age groups. The central panel (central migration, scaled and reallocated) reproduces the primary comparison shown in Figure 4B.

Under the central migration scenario, cumulative admissions for the 2047 counterfactual ranged from 625,859 under the optimistic “scaled by age” deployment (a 30·3% increase over the baseline of 480,492) to 1,193,883 under the conservative “historic vaccinations” assumption (a 148·5% increase), a range more than twenty times wider than that attributable to migration uncertainty alone. The “historic vaccinations” scenario produces a particularly stark result: even under the low migration assumption, the 2047 counterfactual under this strategy results in more than twice the cumulative admissions of the historic baseline. Taken together, these results indicate that future pandemic burden in an older population is highly sensitive to the strategy by which vaccine supply is scaled and allocated across age groups, and largely robust to uncertainty in future migration trends.

## Discussion

Using an age-structured transmission model previously calibrated against the real trajectory of the COVID-19 pandemic in England and used to inform UK government policy,^24^ we show that an equivalent pandemic occurring under the ONS principal 2047 population projection for England would result in 674,004 cumulative hospital admissions and 164,977 hospital deaths up to 24 February 2022 — increases of 40·3% and 49·4% respectively over the historic baseline, despite the underlying population being only 13·9% larger. Despite these higher totals, the underlying basic reproduction number was not significantly impacted by the demographic shift. This disproportionate scaling indicates that the consequences of population ageing for pandemic burden cannot be inferred from demographic growth alone. Critically, our sensitivity analysis shows that while the magnitude of this excess is relatively insensitive to uncertainty in future migration numbers, it is highly sensitive to how vaccine supply is scaled and allocated across age groups. Under our central vaccine deployment assumption, in which total daily doses scale with overall population growth and reallocated following JCVI age-priority order, cumulative admissions under the 2047 counterfactual were 40·3% higher than baseline. Under an optimistic scenario in which doses to each age group track that group’s own population growth, this excess was substantially attenuated (30·3%), while under a conservative assumption of unchanged absolute vaccine supply, admissions were 148·5% higher than baseline — more than double. The implication is stark: the burden associated with an equivalent future pandemic in an older England is not fixed by demography, but is strongly modifiable through preparedness of vaccine supply and deployment infrastructure.

It is important to be clear about what this study does and does not claim. We do not purport to predict the nature or burden of any future pandemic. The next pandemic will differ from COVID-19 in its pathogen, its mode of transmission, its age-specific severity profile, the speed and availability of countermeasures, and the policy environment in which it emerges. Any such factors could substantially amplify, attenuate, or qualitatively alter the findings we describe. Rather, we exploit the unprecedented richness of age-stratified epidemiological, clinical, and vaccination data generated during the COVID-19 pandemic to ask a deliberately controlled question: holding every aspect of the pandemic constant except population age structure, how much does that single change in demographic composition matter? The COVID-19 pandemic is the first outbreak for which sufficient age-specific data exist at the population level to permit this kind of analysis. By using a model validated across dozens of data streams in seven NHS regions simultaneously, we can isolate the effect of demographic composition from measurement noise in a way that cruder burden-estimation approaches cannot. Our findings should therefore be read as a quantification of the structural vulnerability that population ageing creates.

Our results are notably robust to uncertainty in future migration trends. Across low, central, and high ONS migration scenarios under the central vaccine deployment assumption, the range of simulated cumulative hospital admissions for the 2047 counterfactual spans only 664,451 to 689,833, a spread of less than 4%. This robustness persists across the 2027 and 2037 intermediate population projections as well. Migration, while consequential for total population size (with low and high scenarios differing by 4·2 million fewer and 5·4 million more people respectively by 2047), drives relatively modest changes in the age structure of the population compared to the dominant demographic trends of declining fertility and rising life expectancy (see Section 5.1 of Supplementary Appendix). This is unsurprising – the small long-term impact of migration on age structures of migrant-receiving developed countries has been well documented in multiple simulation studies, also including the United Kingdom.^25^ In our case, it is precisely the age structure, not total population size, that drives the mechanism we identify.

These findings have profound practical implications: they suggest that uncertainty about future migration, a subject of considerable political and societal debate in England, contributes relatively little additional uncertainty to pandemic preparedness planning. The key demographic inputs to such planning - the projected size of the oldest age cohorts and the resulting demand on a JCVI-style priority vaccine queue - are largely stable across different migration scenarios.

Our work sits within a small but growing literature on the infectious disease consequences of demographic change. Early theoretical work demonstrated that declining fertility in Italy was reshaping the age distribution of measles incidence and morbidity, with population ageing predicted to shift cases towards older, higher-risk adults even in the presence of sub-optimal vaccination.^26^ Subsequent modelling in Australia showed that demographic change modifies both age- and household-level disease dynamics and can cause models that assume static population structure to overestimate the impact of vaccination programmes.^27^ Burden-of-disease analyses in the Netherlands demonstrated that incorporating realistic population ageing dynamics, rather than assuming a static demographic structure, predicted a growing divergence from static-model estimates over the projection horizon: dynamic modelling predicted disease burden up to 2.3-fold and 1.5-fold higher than a static demography for seasonal influenza and hepatitis B respectively by 2030.^28^ A related analysis of hepatitis A similarly found higher burden under assumptions of demographic change, though here population ageing was the smaller of two compounding drivers, alongside the replacement of naturally-immune birth cohorts by susceptible younger cohorts.^29^

More recently, a microsimulation of the Belgian population found that demographic ageing was associated with smaller overall attack rates for COVID-19 and influenza outbreaks but a substantially larger burden of mortality,^30^ in accordance with our findings. Our study both extends and departs from this prior literature in important respects. Methodologically, we are the first to combine a pandemic transmission model validated against real surveillance data with official long-range demographic projections and a reconstruction of the real-world vaccine deployment policy, enabling attribution of excess burden to demographic change rather than any co-varying factor. Substantively, we identify the mechanism of interaction between a growing elderly priority cohort and a fixed-order vaccine allocation queue, that has not previously been described, and which points to a specific, addressable policy vulnerability rather than a predetermined consequence of population ageing.

Several limitations of our analysis warrant further discussion. First, our model is stratified by age but does not explicitly represent households or long-term care facilities. Prior work has highlighted that clustering of the oldest and most clinically vulnerable individuals within care settings can be a decisive driver of the mortality burden of respiratory infections in ageing populations.^30^ As the 80+ population grows by approximately 85% under the 2047 projection, the care-home population will grow commensurately, and the transmission dynamics within those settings, characterised by high contact density, shared living spaces, and staff rotation, are not captured in our model. Our estimates may therefore understate the concentration of mortality burden within such facilities.

Second, we held the timing and stringency of all non-pharmaceutical interventions fixed at their historical values across both scenarios, as well as dates of variant emergence. In reality, a government observing hospital admissions rising as dramatically as projected in July 2021 of the central counterfactual (Figure 4A) would likely have intervened more stringently, dampening the effect we describe. Our estimates should therefore be understood as the demographic effect under an unchanged policy response, isolating the demographic effect from adaptive policy behaviour. Finally, though re-scaled for reciprocity, we applied an age-specific contact matrix derived from the same pre-pandemic UK contact data.^9^ Contact patterns are themselves subject to demographic influence: as populations age, the relative contributions of different settings and age-group pairings to overall mixing shift, and these patterns are not universal. Systematic reviews have shown that the decline in social contacts with advancing age, while consistent across high-income settings, varies substantially across countries of different income levels.^10^ It remains to be seen how contact behaviours may continue to evolve in the coming decades.

Population ageing is among the most predictable features of England’s demographic future, and yet its consequences for pandemic preparedness have received comparatively little attention. Our findings give quantitative substance to that concern. A pandemic with broadly COVID-19-like characteristics, occurring in 2047, would place demands on England’s health system substantially in excess of those that would be predicted by projecting today’s burden forward in proportion to population growth alone. Those demands would be mediated through the mechanics of how vaccines are allocated across age groups under time pressure and supply contraints. This vulnerability is not inevitable: our results show that a demographically-adaptive vaccine deployment strategy, designed in advance to match supply to the projected age distribution of demand, can substantially attenuate the excess. Such a strategy requires revising existing deployment frameworks against current demographic projections before the next pandemic begins, rather than after it has started. Investment in the manufacturing, procurement, and logistical infrastructure needed to deliver such a strategy, particularly the front-loaded supply surge required to protect a much larger oldest-age cohort rapidly, may represent one of the highest-value, most actionable steps England and comparable countries can take now to reduce the pandemic risk their own demographic trajectories are already generating.

## Data Availability

All data and source code produced are available online at https://github.com/thomrawson/sarscov2-pandemic-ageing

## Data Sharing

All data files and source code required to reproduce this analysis are publicly available at GitHub.

## Contributors

TR conceived the study, analysed the data, and created the figures. TR, DV, JB, JBD, and MCM wrote the original draft of the manuscript. TR, PNPG, ESK, DV, KAG, LKW, WH, RS, RGF, AC, and NMF developed the code base. All authors contributed to interpretation, investigation, and reviewing and editing of the manuscript. PNPG, WH, and KAMG, accessed and verified the underlying data used in the study.

## Acknowledgments

TR, DV, JB, JBD and MCM are supported by the Leverhulme Trust Large Centre Grant RC-2018-003 for the Leverhulme Centre for Demographic Science. MCM is supported by Einstein Foundation Berlin (EZ-2019-555-2) and ESRC/UKRI Connecting Generations (ES/W002116/1). TR is supported by the Moh Family Foundation. PNPG, ESK, CM, KAMG, LKW, WH, RS, AC and NMF acknowledge funding from the MRC Centre for Global Infectious Disease Analysis (reference MR/X020258/1), funded by the UK Medical Research Council (MRC). ESK, LKW, AC, and NMF acknowledge funding from the National Institute for Health Research (NIHR) Health Protection Research Unit in Health Analytics & Modelling (NIHR207404), a partnership between the UKHSA, London School of Hygiene & Tropical Medicine and Imperial College of Science, Technology, & Medicine. PNPG acknowledges funding from Community Jameel through the Abdul Latif Jameel Institute for Disease and Emergency Analytics at Imperial College London. During the course of the study - JBD received funding from the European Research Council (ERC-2021-CoG-101002587); CM received funding from Schmidt Sciences (6-22-63345); KAMG received funding from Gavi (226727_Z_22_Z), BMGF (INV-034281 and INV-009125/OPP1157270) and the Wellcome Trust via the Vaccine Impact Modelling Consortium.

The funders had no involvement in the study design; in the analysis; in the writing of the paper; or the decision to submit the paper for publication.

